# Mean Dynamics Index: a useful tool to identify motor psychogenic non epileptic seizures

**DOI:** 10.1101/2024.02.07.24302379

**Authors:** Raz Winer, Shahram Shlomo Shahkoohi, Moshe Herskovitz

**Author notes:** **Corresponding address:** Moshe Herskovitz, Epilepsy Service, Dept of Neurology, Rambam Medical Center, Technion Faculty of Medicine, 1 Efron St. Haifa, Israel 31096. **Authors’ contribution:** Raz Winer MD- Analysis and interpretation of data, writing and revision of the manuscript. Shahram Shlomo Shahkoohi MD- Data acquisition Moshe Herskovitz MD – Conceptualization of the study, Analysis and interpretation of data and revision of the manuscript.

## Abstract

Timely and accurate diagnosis of Psychogenic Non-Epileptic Seizures (PNES) is crucial. Aside from potential diagnostic delays, patients with PNES often undergo unnecessary pharmacological or invasive treatments. Presently, effective bedside tools for distinguishing PNES from epileptic seizures (ES) remain elusive, and the ‘gold standard’ diagnosis relies primarily on patient history and prolonged video EEG monitoring. In this study, we developed a simple clinical tool - the Mean Dynamic Index (MD) - to differentiate PNES from ES.

We divided the body into five anatomical regions: the head and face, two upper extremities, and two lower extremities. Due to limited movement potential, the trunk was excluded from consideration. Among these five areas, only actively involved regions were considered in the score calculation. Each distinct motor feature observed contributed a point to the regional summation, with each region’s score comprising the Regional Dynamic Index (RDI). The Mean Dynamic Index (MDI) represents the average of all RDIs.

Sixty consecutive patients admitted to the VEEG monitoring unit were evaluated. Of these, 15 patients presented primarily with motor symptoms. Eight were diagnosed with PNES, while seven had epileptic seizures. The mean MDI was 1.2±0.4 in the PNES group and 2.8±0.77 in the ES group (p<0.001). The mean MDI/duration ratio was 0.31±0.38 for PNES and 3.5±2.4 for ES (p < 0.003). An MDI score of 1.665 yielded a specificity of 87.5% and sensitivity of 100% for diagnosing ES. A low MDI score (<1.66) in motor seizures indicates limited variability in the movement profile of each body part, suggesting a PNES etiology. Additionally, as the MDI/time ratio decreases, PNES becomes more likely. Furthermore, we observed that an RDI of 3 or higher completely differentiated between PNES and ES. These findings offer a valuable bedside tool for distinguishing between the two conditions.

## Background

Ensuring a reliable and timely diagnosis of Psychogenic Non-Epileptic Seizures (PNES) is paramount. In addition to potential delays in diagnosis, these patients often undergo unnecessary pharmacological or invasive treatments [1]. Presently, efficient bedside tools for distinguishing PNES from epileptic seizures (ES) have not been identified, and the current ‘gold standard’ diagnosis relies primarily on patient history and prolonged video EEG monitoring[2].

Epileptic seizures (ES) arise from the propagation of epileptic discharges throughout the epileptic network, activating various functional areas. For instance, the Jacksonian march, characterized by clonic movements spreading from distal to proximal limb areas, exemplifies this phenomenon in central lobe epilepsy[3]. Similarly, alterations in flexion-extension of tonic posture in mesial frontal lobe epilepsy and changes in dystonic limb position in temporal lobe epilepsy represent well-known epileptic patterns reflecting the evolution of seizures over time[4].

The precise pathophysiology of PNES remains unclear, despite mounting evidence of brain connectivity abnormalities in these patients[5-7]. A resting-state fMRI study employing graph theory among twenty-three PNES patients and twenty-five healthy controls revealed greater nodal degrees (indicating hyper-connectivity) in the right caudate, left orbital part of the left inferior frontal gyrus, and right paracentral lobule. Conversely, lesser nodal degrees (indicating hypoconnectivity) were observed in the right insula, right putamen, and right middle occipital gyrus. Researchers suggested that hypo-connectivity areas may be implicated in emotion processing and movement regulation, while hyper-connectivity areas may play a role in inhibiting unwanted movements and cognitive processes[8].

However, it is well-established that PNES does not stem from abnormal electrical discharges[9]. Moreover, PNES typically exhibits a relatively recurrent and consistent phenotype. Numerous studies have demonstrated this stereotypic tendency, characterized by the recurrence of the same typical motor behavior in individual patient events[10-12]. In a previous study conducted by our group, we illustrated that different PNES events in one patient manifest similar characteristics, including the sequence of movements, the predominant region involved, and the frequency of clonic-like movements[13]. Despite PNES being perceived as a dramatic event, the motor behavior in each involved region remains stable and simplistic.

Accordingly, we hypothesized that PNES patients would exhibit fewer motor alternations than ES patients during motor-predominant events.

## Methods

This retrospective study aimed to reflect our observation that Psychogenic Non-Epileptic Seizures (PNES), unlike Epileptic Seizures (ES), exhibit a consistent, repetitive, and recurrent nature. To operationalize this concept, we devised the Mean Dynamic Index (MDI), a scoring system that quantifies the occurrence of unique motor patterns during motor-predominant seizures.

We segmented the body into five anatomical regions: the head and face, two upper extremities, and two lower extremities. Owing to limited movement potential, the trunk was excluded from consideration. Only actively involved areas among these five regions contributed to the score calculation. Each distinct motor feature observed added a point to the regional summation, thereby constituting the Regional Dynamic Index (RDI). The Mean Dynamic Index (MDI) represents the average of all RDIs.

For instance, a seizure exhibiting distal hand clonus progressing to proximal hand clonus would earn an RDI of 2 (1 point for distal clonus and another for proximal clonus), resulting in an MDI of 2 if this were the sole area involved. Conversely, an attack characterized by waxing and waning distal right-hand clonus would earn an RDI of 1 for the right upper extremity. A high MDI score indicates a greater number of unique motor patterns observed per region during the seizure.

To achieve 80% power and 5% significance, assuming MDI score differences of at least 25% between groups, the sample size was calculated. An independent evaluator, a medical student (‘the first evaluator’), reviewed medical records of patients admitted to the video-EEG monitoring unit at Rambam Medical Center between November 1st, 2015, and May 31st, 2016. Twenty consecutive patients with established diagnoses of motor PNES and motor ES were included (10 in each group). The first evaluator extracted the video component of the first seizure of each patient and provided them to two blinded evaluators (‘evaluator two,’ a neurologist, and ‘evaluator three,’ a trained epileptologist).

Each evaluator calculated the MDI for every event, with the final MDI being the average score of the two evaluators. These evaluators were blinded to additional data, including diagnosis, medical records, EEG recordings, or other evaluator scores. All seizures were analyzed from start to finish.

Additionally, we measured the ratio between MDI and seizure duration. Demographic and clinical information, including age, age at onset, gender, number of medications at admission, and present and past anti-seizure medications, was collected.

Descriptive statistics were conducted using SPSS v25 for Windows. Tailored t-tests were employed to compare means between ES and PNES groups. Receiver operating characteristic analysis was performed to determine the MDI score with the highest sensitivity and specificity for differentiating between ES and PNES patients.

All study procedures were approved by Rambam Health Care Campus institutional review board, Helsinki approval number-0283-17-RMB.

## Results

To identify 20 consecutive patients with either motor Epileptic Seizures (ES) or motor Psychogenic Non-Epileptic Seizures (PNES) between November 1st, 2015, and May 31st, 2016, the first evaluator reviewed 60 files. A total of 20 videos (one for each patient) were provided to the second and third evaluators. During video evaluation, five patients were excluded due to technical and semiological reasons: two patients were either obstructed or partially obscured from view in the camera field, and three patients exhibited mainly dialeptic semiology without apparent motor activity. Among the remaining 15 patients, eight were diagnosed with PNES, and seven had epileptic seizures (see Figure 1). The average age of patients in the PNES and ES groups was 29.37±11.2 years and 30.85±12.7 years, respectively, with mean durations from onset to diagnosis of 4.35±5.5 years and 13.48±11.7 years, respectively. The average number of medications currently used was 1.37±1.5 and 3.42±2.1 in the PNES and ES groups, respectively (see Table 1 for patient demographics and clinical features).

**Table 1:** Patients’ demographics and clinical characteristics including.

|  | PNES (n=8) | ES (n=7) | P<br>value |
| --- | --- | --- | --- |
| Age (years) ± SD | 29.37±11.2 | 30.85±12.7 | 0.81 |
| Male (%) | 37.5 | 43 | 0.83 |
| Mean total ASM received ± SD | 2.87±4 | 6.42±3.75 | 0.1 |
| No of Current ASM±SD | 1.37±1.5 | 3.42±2.1 | 0.049 |
| Disease Duration (years) ± SD | 4.3±5.5 | 13.48±11.7 | 0.07 |

**Figure 1:**
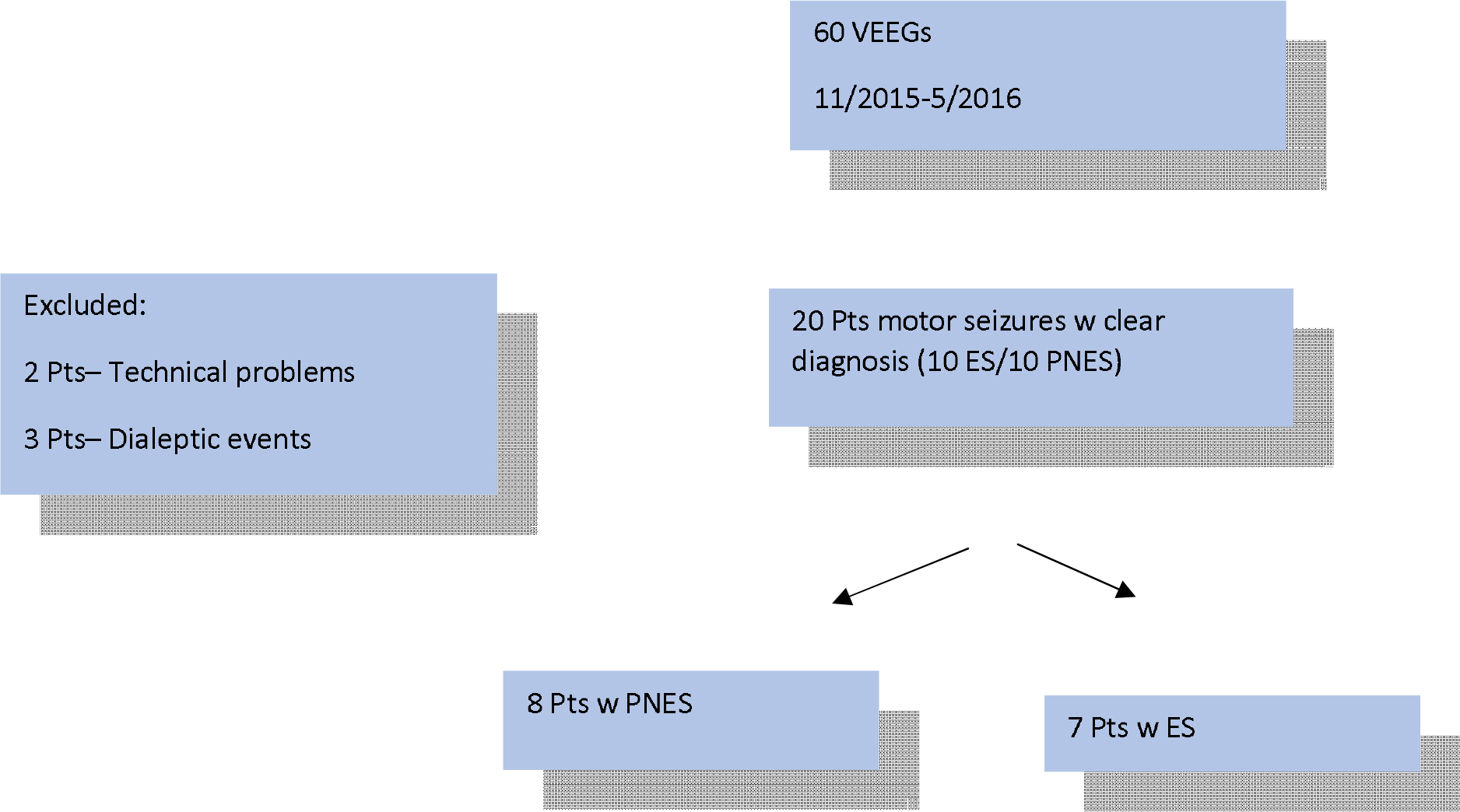
patients enrollment flowchart

The mean Mean Dynamic Index (MDI) was 1.2±0.4 in the PNES group and 2.8±0.77 in the ES group (p<0.001). Furthermore, the mean MDI/duration ratio was 0.31±0.38 and 3.5±2.4 in the PNES and ES groups, respectively (p < 0.003). Figure 2 (A-D) provides a summary of the study’s main results.

**Figure 2:**
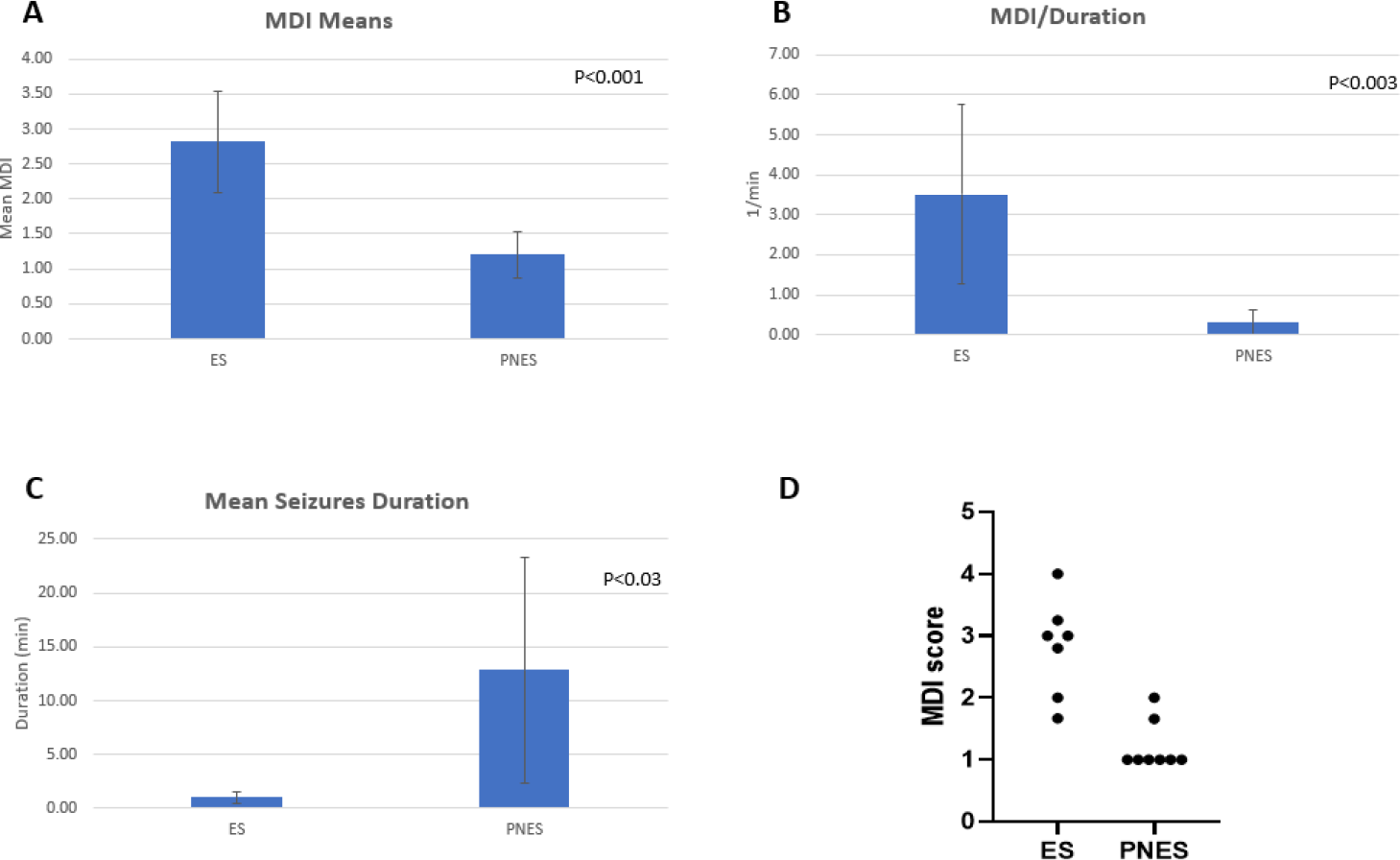
Figure 2A- comparison of mean MDI between ES and PNES groups, 2B- comparison of mean MDI to seizure duration between ES and PNES groups, 2C –comparison of mean seizure duration between ES and PNES groups, 2D- scattered plot of all patients’ MDI

To determine the MDI with the highest sensitivity and specificity for distinguishing between ES and PNES groups, a Receiver Operating Characteristic (ROC) curve was plotted (see Figure 3). An MDI score of 1.33 yielded a specificity of 75% and sensitivity of 100% for diagnosing ES, while an MDI score of 1.665 resulted in a specificity of 87.5% and sensitivity of 100% for ES diagnosis.

**Figure 3:**
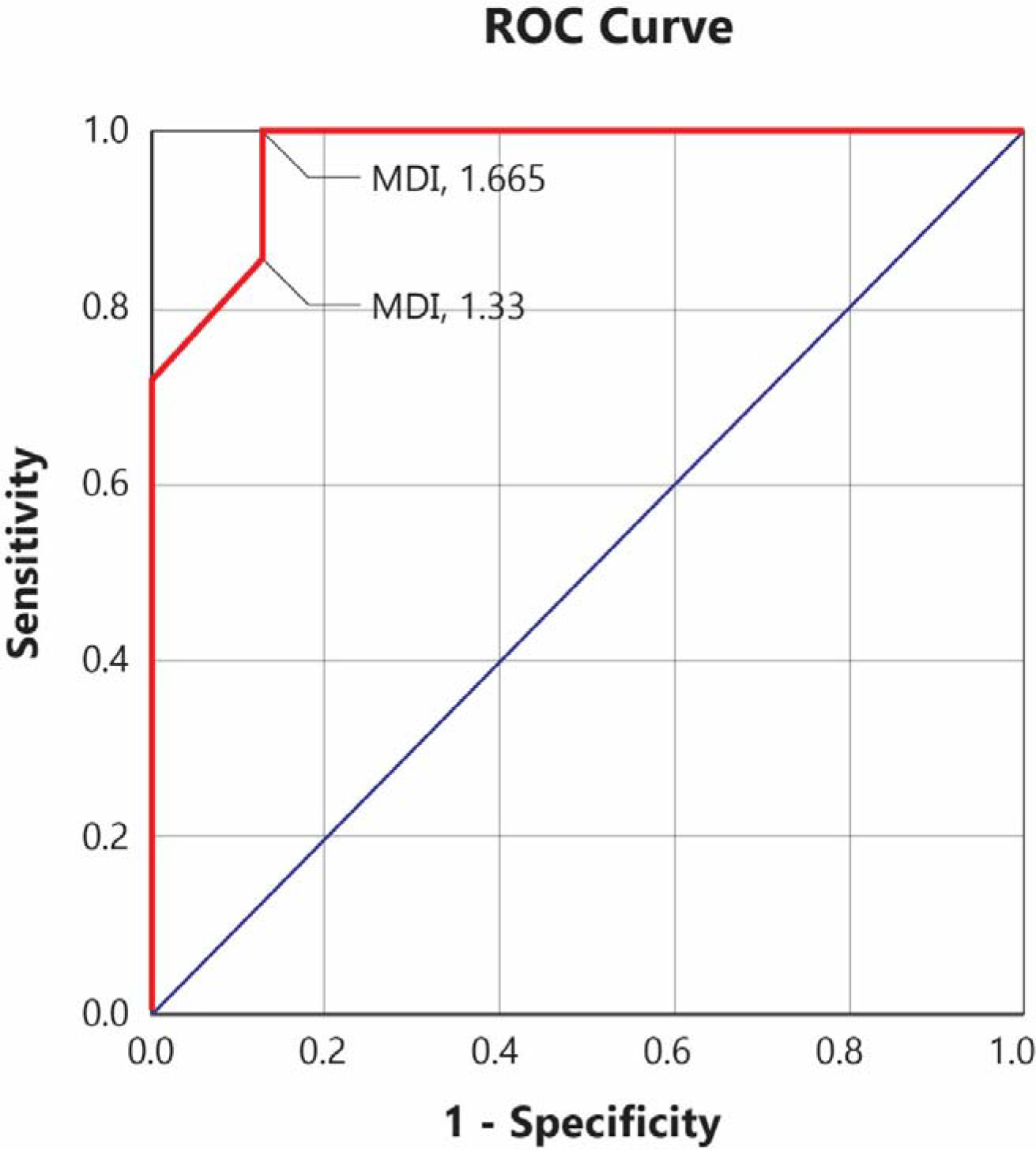

Table 2 outlines the components of the MDI score, indicating that an RDI of 3 and above completely differentiates between ES and PNES (The patients are assigned a serial number for coding purposes, ensuring their identities remain confidential and cannot be discerned).

**Table 2:** MDI score components.

| Patient No | Diagnosis | MDI score | Duration (minutes) | MDI/Duration ratio | RDI Indexes |
| --- | --- | --- | --- | --- | --- |
| 1 | PNES | 1 | 40 | 0.025 | Head and Face – 1<br>Left upper extremity – 1<br>Right upper extremity – 1 |
| 2 | ES | 3 | 1 | 3 | Head and Face – 4<br>Left upper extremity – 4<br>Right upper extremity – 3<br>Left lower extremity – 2<br>Right lower extremity – 2 |
| 3 | PNES | 1.66 | 16 | 0.10375 | Head and Face – 2<br>Left upper extremity – 1<br>Right upper extremity – 1 |
| 4 | PNES | 1 | 1 | 1 | Head and Face – 1<br>Left upper extremity – 1 |
|  |  |  |  |  | Right upper extremity – 1 |
| 5 | PNES | 1 | 6 | 0.166667 | Head and Face – 1 |
| 6 | ES | 2.8 | 0.5 | 5.6 | Head and Face – 4<br>Left upper extremity – 3<br>Right upper extremity – 2<br>Left lower extremity – 2<br>Right lower extremity – 3 |
| 7 | ES | 2 | 0.25 | 8 | Head and Face – 2<br>Left upper extremity – 1<br>Right upper extremity – 1<br>Right lower extremity – 4 |
| 8 | ES | 3.25 | 1.166 | 2.787307 | Left upper extremity – 3<br>Right upper extremity – 4<br>Left lower extremity – 3<br>Right lower extremity – 3 |
| 9 | ES | 4 | 2 | 2 | Head and Face – 4<br>Single extremity – 4 |
| 10 | PNES | 1 | 1.2 | 0.833333 | Left upper extremity – 1 |
| 11 | PNES | 1 | 13 | 0.076923 | Head and Face – 1<br>Left upper extremity – 1<br>Left lower extremity – 1 |
| 12 | PNES | 1 | 17 | 0.058824 | Left lower extremity – 1<br>Right lower extremity – 1 |
| 13 | ES | 1.67 | 0.616667 | 2.708108 | Head and Face – 3<br>Left upper extremity – 2<br>Right lower extremity – 1 |
| 14 | ES | 3 | 1.36 | 2.205882 | Head and Face – 2<br>Left upper extremity – 4<br>Right upper extremity – 3 |
| 15 | PNES | 2 | 8.63 | 0.23175 | Head and Face – 2<br>Left upper extremity – 2 |

## Discussion

This is a proof-of-concept study. In this small cohort of patients, the mean MDI index was 2.8 in the ES group, compared with 1.2 in the PNES group. (p<0.001). MDI/duration ratio was 3.76 and 0.31 in the ES group and PNES group, respectively (p<0.003). Both MDI and MDI/duration measures were significantly different between the two groups.

The former result - a significant difference in the MDI measures - has substantial clinical importance; The MDI rating system can be easily learned, thus enabling accurate diagnosis by a novice healthcare giver when observing a seizure.

Upon further examination of the Regional Dynamic Index (RDI) scores, it was determined that a definitive cutoff can indeed be established to distinguish between all PNES and ES cases within our dataset with complete accuracy. The maximal RDI score within the PNES group did not exceed 2, while the minimal RDI score observed within the ES group was strictly greater than 2. Consequently, an RDI cutoff set at 2 serves as an unequivocal threshold—scores of 2 or below are indicative of PNES, while scores above 2 are predictive of ES. This clear demarcation allowed for 100% differentiation between the two conditions, with no overlap between the groups, thus confirming the robustness of the maximal RDI score as a discriminative measure in this study.

There are two clinical scenarios in which a first responder must determine whether a convulsion represents ES or PNES, and consequently, conclude what should be the next therapeutic step. The first is the bed-side scenario - when a seizure is seen in real-time by a healthcare professional. The second occurs when a caregiver or the patient himself brings a movie-clip that documents a past event.

Previous studies have shown that experienced epileptologists can differentiate between motor-predominant ES and PNES by observing video recordings as-accurately-as by analyzing a full video EEG monitoring[14]. Another study showed differences in the accuracy of diagnosis given by different epilepsy specialists based on video recording alone.

Nonetheless, some of them were no less accurate than the epileptologist who read the whole medical chart and the video-EEG exam in motor-predominant seizures.[15] Several groups used machine learning algorithms for frequencies analysis to classify PNES VS other event-types, utilizing variable devices to perform the acquisition of movement frequencies[16–20]. The researchers attributed this approach’s accuracy to the inherent variability of ictal discharge, as opposed to the non-epileptic, and thus consistent, movement rhythmicity during PNES. Other studies supported this hypothesis: One study showed variability in the EEG’s predominant frequency throughout an epileptic seizure, whereas the frequency range during PNES remained narrow[16]. Further studies revealed differences in time-frequency mapping of the rhythmic limb movements between PNES and ES groups[17].

Articles dealing with the difference between initiation, propagation, and termination have shown the fundamental mechanism to be various dynamic network expressions and interactions. This mechanism is expressed in different rhythms or frequencies throughout the seizure and from a clinical perspective – in spatiotemporal evolution during a seizure[21–23]. We believe these essential rhythms and spreading dynamics might explain the intuitive PNES\ES differentiation ability of experienced epileptologists: subconsciously, they look for the variability of frequencies, local spread and change in each area, which characterizes ES and lacks in PNES. These expected patterns - of simultaneously evolving rhythms and spatial spread - constitute the hallmark semiology of many well-known epilepsy types: e.g., the consecutive appearance of eye deviation, ictal sound, and Chapo-Jun-Darm (all belong to the “head and face region’”) in frontal lobe epilepsy ; dystonia, automatic movements in upper extremities involved in temporal lobe seizures, etc[3,4].

The MDI scoring method was conceptualized to reflect this dual variability - both in frequencies and in local spatial evolvement - over time. However, instead of device-dependent frequency acquisition (e.g., accelerometer or EMG wristband[17–20], EEG recordings analysis[16], etc.), we use an easy-to-learn method to quantify the number of observed changes in the involved region. Simply, any change that seems like a “new occurrence” in the eyes of the beholder equals a point. MDI is the average of all areas scores, and in a way, it quantifies the amount in which the convulsion went through onset-propagation-termination.

Albeit a small cohort, we established a statistically significant difference in the MDI score between the ES and PNES groups. Previous research of our group demonstrated relative uniformity of seizures semiology in a single PNES patient. We decided to include only one episode of each patient in the analysis to avoid overfitting.

Moreover, there was a trend toward MDI-score-cutoff (MDI>1.66) between the two groups. An MDI-scoring-cutoff, if established in more extensive studies, has substantial clinical importance. It will facilitate bed-side decision making in case of PNES\ES deliberation and, consequently, avoid unwanted interventions, admissions, or polypharmacy repercussions. Previous studies have suggested numerous possible characteristics of PNES. Suggested features of PNES semiology include eyes-closing, shaking the head from side to side, pelvic thrust, ictal crying, and more[24–29]. Some “time-dependent” PNES characteristics have also been suggested, including long-duration, fluctuating presentation, occurrence during pseudo-sleep, and others[15,28–31]. None of these features has been established as specific and sensitive enough to utilize as a bedside tool for decision making.

Many studies address biomarkers that can differentiate PNES from ES. In a study that investigated whether anion gap (AG), bicarbonate, and the Denver Seizure Score (DSS) could differentiate between PNES and ES, AG was associated with ES (OR of 1.733, 95% confidence interval = 1.24–2.42)[32]. Another study found maximal-ictal and postictal HR to be useful in differentiating bilateral tonic-clonic ES from PNES[33]. In a study addressing the verbal report of an event by the patient, the researchers succeeded in classifying PNES and ES groups based on language analysis of an interview following the event[34].

Until now, no sign, feature, or biomarker has proved the ability to become a bedside tool for early diagnosis of PNES.

A possible interpretation for the low MDI scores among PNES patients in our study might be a production of repetitive movements during dissociation. Altered consciousness is present in many PNES and may result from a process of dissociation[35]). An SEEG study that recorded two patients’ PNES events demonstrated a global decrease of Frontal connectivity, particularly prominent in connections involving the anterior insula and parietal cortex[36]. The researchers interpreted these findings as a functional disconnection, resulting from alterations in long-distance Fc involving the parietal cortex, and a disconnection of the anterior insula (AI), that cause lack of normal continuous production of predictions, thus contributing to a feeling of being “disconnected”[36].

Previous studies also suggested a correlation between decreased interictal Fc and PNES: few EEG studies demonstrated decreased inter-ictal Fc between frontal and posterior regions[37] or intra-regional[38]. fMRI studies found decreased Fc in several locations, including the parietal region[39–42]. A PET study demonstrated interictal focal hypometabolism, particularly in the right parietal area[43]. Other previous studies supported the insular cortex’s role in the conscious representation of self [44], interception, and emotions[45]. the researchers hypothesized that this “disconnection” might serve as a protective mechanism to avoid representations that would be considered threatening[36]. A diagnostic delay of PNES has a dual negative effect: first, the admission of unnecessary medication and interventions, including multiple AEDs, ICU admissions, mechanical ventilation, and possible iatrogenic complications. Moreover, these unwanted interventions might even worsen the PNES [1]. Secondly, a good prognosis of PNES depends on the early start of psychological interventions such as CBT and psychotherapy, especially in the presence of sexual or child abuse in history, a common situation in PNES patients, up to 88% in some series [31]. However, no tool currently exists to shorten the average delay of 7-9 years from onset to diagnosis[1].

Our study has several limitations: First, the number of events was small. Indeed, we have decided to include a single event per patient due to the expected events-uniformity in a specific patient, both in PNES and ES [13], and to exclude recent video recordings in order to have no recollection of interpretations made by us. However, these two limitations, conducted to increase validity - took the price of low event number. Secondly, a trained epileptologist (examiner1) and neurologist (examiner 2) calculated the MDI scores; they might have used their pre-existing intuition without being aware of that. However, only a naive observer classification, made after teaching him the MDI method, could prove its clinical importance and usefulness. Further research is needed in which the MDI method is explained to possible non-epileptic first responders, and the contribution to accurate classification of ES\PNEs is measured. It is also essential to address the unique entity of EPC, which might be misclassified as PNES based on MDI scoring alone. However, these are not common, easy to investigate by EEG due to their continuity, have a distinct presentation, and do not mimic GTCS or other event types that allegedly require status-epilepticus workup. It is also important to note that correct MDI scoring requires full-length and full-body observation of the seizure and cannot be conducted based on partial video recording or by observing only part of the seizure.

In conclusion - VEEG is currently the gold-standard tool for diagnosing PNES [2]; However, frequently unavailable. For this reason and others, the average diagnostic delay is 7-9 years [1]. Throughout that period, the patient might accumulate iatrogenic complications and the repercussion of avoiding emotional treatment. Previous studies established the ability of experienced Epilepsy specialists to diagnose motor PNES as accurate as VEEG, based on observation only. However, until now, no bedside or fast tool to aid this decision was established.

We suggest the MDI index, which allows an immediate assessment and PNES\ES classification of observed seizure. We believe that the MDI index, namely - the non-variability of regional involvement - might underlie the well-established ability of experienced epileptologists to differentiate motor ES from PNES with great accuracy. Furthermore, although it was not the primary focus of our study, a maximal RDI of 3 or higher can completely differentiate between ES and PNES.

The MDI scoring system is a simple tool to learn and can significantly improve healthcare professionals’ clinical decision-making regarding PNES diagnosis and treatment decisions, thus spare unnecessary, costly, and harmful interventions.

## Data Availability

All data produced in the present study are available upon reasonable request to the authors

## References

[1] Asadi-Pooya AA, Sperling MR. Epidemiology of psychogenic nonepileptic seizures. Epilepsy Behav 2015;46:60–5. 10.1016/j.yebeh.2015.03.015.

[2] Kanemoto K, LaFrance WC, Duncan R, Gigineishvili D, Park SP, Tadokoro Y, et al. PNES around the world: Where we are now and how we can close the diagnosis and treatment gaps—an ILAE PNES Task Force report. Epilepsia Open 2017;2:307–16. 10.1002/epi4.12060.

[3] Foldvary-Schaefer N, Unnwongse K. Localizing and lateralizing features of auras and seizures. Epilepsy Behav 2011;20:160–6. 10.1016/j.yebeh.2010.08.034.

[4] Stoyke C, Bilgin Ö, Noachtar S. Video atlas of lateralising and localising seizure phenomena. Epileptic Disord 2011;13:113–24. 10.1684/epd.2011.0433.

[5] Asadi-Pooya AA. Neurobiological origin of psychogenic nonepileptic seizures: A review of imaging studies. Epilepsy Behav 2015;52:256–9. 10.1016/j.yebeh.2015.09.020.

[6] Allendorfer JB, Nenert R, Hernando KA, DeWolfe JL, Pati S, Thomas AE, et al. FMRI response to acute psychological stress differentiates patients with psychogenic non-epileptic seizures from healthy controls – A biochemical and neuroimaging biomarker study. NeuroImage Clin 2019;24:101967. 10.1016/j.nicl.2019.101967.

[7] Foroughi AA, Nazeri M, Asadi-Pooya AA. Brain connectivity abnormalities in patients with functional (psychogenic nonepileptic) seizures: A systematic review. Seizure 2020;81:269–75. 10.1016/j.seizure.2020.08.024.

[8] Amiri S, Mirbagheri MM, Asadi-Pooya AA, Badragheh F, Ajam Zibadi H, Arbabi M. Brain functional connectivity in individuals with psychogenic nonepileptic seizures (PNES): An application of graph theory. Epilepsy Behav 2021;114:107565. 10.1016/j.yebeh.2020.107565.

[9] Bodde NMG, Brooks JL, Baker GA, Boon PAJM, Hendriksen JGM, Aldenkamp AP. Psychogenic non-epileptic seizures-Diagnostic issues: A critical review. Clin Neurol Neurosurg 2009;111:1–9. 10.1016/j.clineuro.2008.09.028.

[10] Seneviratne U, Reutens D, D’Souza W. Stereotypy of psychogenic nonepileptic seizures: Insights from video-EEG monitoring. Epilepsia 2010;51:1159–68. 10.1111/j.1528-1167.2010.02560.x.

[11] Asadi-Pooya AA, Tinker J, Fletman EW. How variable are psychogenic nonepileptic seizures? A retrospective semiological study. J Neurol Sci 2017;377:85–7. 10.1016/j.jns.2017.03.054.

[12] Vogrig A, Hsiang JC, Ng J, Rolnick J, Cheng J, Parvizi J. A systematic study of stereotypy in epileptic seizures versus psychogenic seizure-like events. Epilepsy Behav 2019;90:172–7. 10.1016/j.yebeh.2018.11.030.

[13] Herskovitz M. Stereotypy of psychogenic nonepileptic seizures. Epilepsy Behav 2017;70:140–4. 10.1016/j.yebeh.2017.02.015.

[14] Tatum WO, Hirsch LJ, Gelfand MA, Acton EK, Lafrance WC, Duckrow RB, et al. Assessment of the Predictive Value of Outpatient Smartphone Videos for Diagnosis of Epileptic Seizures. JAMA Neurol 2020;77:593–600. 10.1001/jamaneurol.2019.4785.

[15] Erba G, Giussani G, Juersivich A, Magaudda A, Chiesa V, Laganà A,. et al. The semiology of psychogenic nonepileptic seizures revisited: Can video alone predict the diagnosis? Preliminary data from a prospective feasibility study. Epilepsia 2016;57:777–85. 10.1111/epi.13351.

[16] Vinton A, Carino J, Vogrin S, MacGregor L, Kilpatrick C, Matkovic Z, et al. “Convulsive” nonepileptic seizures have a characteristic pattern of rhythmic artifact distinguishing them from convulsive epileptic seizures. Epilepsia 2004;45:1344–50. 10.1111/j.0013-9580.2004.04704.x.

[17] Bayly J, Carino J, Petrovski S, Smit M, Fernando DA, Vinton A, et al. Time-frequency mapping of the rhythmic limb movements distinguishes convulsive epileptic from psychogenic nonepileptic seizures. Epilepsia 2013;54:1402–8. 10.1111/epi.12207.

[18] Beniczky S, Conradsen I, Moldovan M, Jennum P, Fabricius M, Benedek K, et al. Quantitative analysis of surface electromyography during epileptic and nonepileptic convulsive seizures. Epilepsia 2014;55:1128–34. 10.1111/epi.12669.

[19] Naganur VD, Kusmakar S, Chen Z, Palaniswami MS, Kwan P, O’Brien TJ. The utility of an automated and ambulatory device for detecting and differentiating epileptic and psychogenic non-epileptic seizures. Epilepsia Open 2019;4:309–17. 10.1002/epi4.12327.

[20] Kusmakar S, Karmakar C, Yan B, Muthuganapathy R, Kwan P, O’Brien TJ, et al. Novel features for capturing temporal variations of rhythmic limb movement to distinguish convulsive epileptic and psychogenic nonepileptic seizures. Epilepsia 2019;60:165–74. 10.1111/epi.14619.

[21] Liou JY, Smith EH, Bateman LM, Bruce SL, McKhann GM, Goodman RR, et al. A model for focal seizure onset, propagation, evolution, and progression. Elife 2020;9. 10.7554/eLife.50927.

[22] Merricks EM, Smith EH, Emerson RG, Bateman LM, McKhann GM, Goodman RR, et al. Neuronal Firing and Waveform Alterations through Ictal Recruitment in Humans. J Neurosci 2021;41:766–79. 10.1523/JNEUROSCI.0417-20.2020.

[23] Sadeh S, Clopath C. Inhibitory stabilization and cortical computation. Nat Rev Neurosci 2021;22:21–37. 10.1038/s41583-020-00390-z.

[24] Geyer JD, Payne TA, Drury I. The value of pelvic thrusting in the diagnosis of seizures and pseudoseizures. Neurology 2000;54:227–9. 10.1212/wnl.54.1.227.

[25] Chung SS, Gerber P, Kirlin KA. Ictal eye closure is a reliable indicator for psychogenic nonepileptic seizures. Neurology 2006;66:1730–1. 10.1212/01.wnl.0000218160.31537.87.

[26] Syed TU, Arozullah AM, Suciu GP, Toub J, Kim H, Dougherty ML, et al. Do observer and self-reports of ictal eye closure predict psychogenic nonepileptic seizures? Epilepsia 2008;49:898–904. 10.1111/j.1528-1167.2007.01456.x.

[27] Avbersek A, Sisodiya S. Does the primary literature provide support for clinical signs used to distinguish psychogenic nonepileptic seizures from epileptic seizures? J Neurol Neurosurg Psychiatry 2010;81:719–25. 10.1136/jnnp.2009.197996.

[28] Syed TU, Lafrance WC, Kahriman ES, Hasan SN, Rajasekaran V, Gulati D, et al. Can semiology predict psychogenic nonepileptic seizures? a prospective study. Ann Neurol 2011;69:997–1004. 10.1002/ana.22345.

[29] Xiang X, Fang J, Guo Y. Differential diagnosis between epileptic seizures and psychogenic nonepileptic seizures based on semiology. Acta Epileptol 2019;1:4–8. 10.1186/s42494-019-0008-4.

[30] Seneviratne U, Minato E, Paul E. How reliable is ictal duration to differentiate psychogenic nonepileptic seizures from epileptic seizures? Epilepsy Behav 2017;66:127–31. 10.1016/j.yebeh.2016.10.024.

[31] Gasparini S, Beghi E, Ferlazzo E, Beghi M, Belcastro V, Biermann KP, et al. Management of psychogenic non-epileptic seizures: a multidisciplinary approach. Eur J Neurol 2019;26:205–15. 10.1111/ene.13818.

[32] Li Y, Matzka L, Maranda L, Weber D. Anion gap can differentiate between psychogenic and epileptic seizures in the emergency setting. Epilepsia 2017;58:e132–5. 10.1111/epi.13840.

[33] Au Yong HM, Minato E, Paul E, Seneviratne U. Can seizure-related heart rate differentiate epileptic from psychogenic nonepileptic seizures? Epilepsy Behav 2020;112:107353. 10.1016/j.yebeh.2020.107353.

[34] Biberon J, de Liège A, de Toffol B, Limousin N, El-Hage W, Florence AM, et al. Differentiating PNES from epileptic seizures using conversational analysis on French patients: A prospective blinded study. Epilepsy Behav 2020;111:107239. 10.1016/j.yebeh.2020.107239.

[35] Reuber M, Kurthen M. Consciousness in non-epileptic attack disorder. Behav Neurol 2011;24:95–106. 10.3233/BEN-2011-0315.

[36] Madec T, Lagarde S, McGonigal A, Arthuis M, Benar CG, Bartolomei F. Transient cortico-cortical disconnection during psychogenic nonepileptic seizures (PNES). Epilepsia 2020;61:e101–6. 10.1111/epi.16623.

[37] Knyazeva MG, Jalili M, Frackowiak RS, Rossetti AO. Psychogenic seizures and frontal disconnection: EEG synchronisation study. J Neurol Neurosurg Psychiatry 2011;82:505–11. 10.1136/jnnp.2010.224873.

[38] Barzegaran E, Joudaki A, Jalili M, Rossetti AO, Frackowiak RS, Knyazeva MG. Properties of functional brain networks correlate frequency of psychogenic non-epileptic seizures. Front Hum Neurosci 2012;6:1–13. 10.3389/fnhum.2012.00335.

[39] Van Der Kruijs SJM, Bodde NMG, Vaessen MJ, Lazeron RHC, Vonck K, Boon P, et al. Functional connectivity of dissociation in patients with psychogenic non-epileptic seizures. J Neurol Neurosurg Psychiatry 2012;83:239–47. 10.1136/jnnp-2011-300776.

[40] Ding JR, An D, Liao W, Li J, Wu GR, Xu Q, et al. Altered Functional and Structural Connectivity Networks in Psychogenic Non-Epileptic Seizures. PLoS One 2013;8. 10.1371/journal.pone.0063850.

[41] van der Kruijs SJM, Jagannathan SR, Bodde NMG, Besseling RMH, Lazeron RHC, Vonck KEJ, et al. Resting-state networks and dissociation in psychogenic non-epileptic seizures. J Psychiatr Res 2014;54:126–33. 10.1016/j.jpsychires.2014.03.010.

[42] Li R, Li Y, An D, Gong Q, Zhou D, Chen H. Altered regional activity and inter-regional functional connectivity in psychogenic non-epileptic seizures. Sci Rep 2015;5:1–12. 10.1038/srep11635.

[43] Arthuis M, Micoulaud-Franchi JA, Bartolomei F, McGonigal A, Guedj E. Resting cortical PET metabolic changes in psychogenic non-epileptic seizures (PNES). J Neurol Neurosurg Psychiatry 2015;86:1106–12. 10.1136/jnnp-2014-309390.

[44] Critchley HD, Wiens S, Rotshtein P, Öhman A, Dolan RJ. Neural systems supporting interoceptive awareness. Nat Neurosci 2004;7:189–95. 10.1038/nn1176.

[45] Picard F, Craig AD. Ecstatic epileptic seizures: A potential window on the neural basis for human self-awareness. Epilepsy Behav 2009;16:539–46. 10.1016/j.yebeh.2009.09.013.

